# Involvement of Gut Microbiota in the Development of Psoriasis Vulgaris

**DOI:** 10.1101/2020.11.16.20232025

**Authors:** Chaonan Sun, Ling Chen, Huan Yang, Hongjiang Sun, Zhen Xie, Bei Zhao, Xuemei Jiang, Bi Qin, Zhu Shen

## Abstract

**Background:** Psoriasis is a common chronic recurrent dermatitis. Accumulating observations show gut microbiota dysbiosis in psoriasis.

**Objectives:** We intend to further investigate the relationship between intestinal microbiota and psoriasis development.

**Methods:** We first performed an epidemiological investigation on differences of gastrointestinal discomfort symptoms between psoriatic patients and general population. Then variation of gut microbiota in psoriatic patients (un)treated with Acitretin was analyzed by 16S rRNA sequencing. We last compared recovery status and vital cytokines of mouse psoriasiform models, which were transplanted with fecal microbiota from psoriatic patients or healthy controls.

**Results:** (1) 85.53% of psoriatic patients versus 58.08% of healthy controls presented with at least one gastrointestinal symptom. The prevalence of investigated symptoms (e.g. abdominal distension, constipation) were significantly higher in patients, compared with controls (p<0.05). Increased fart and constipation were significantly correlated with psoriasis (p<0.05, respectively). (2) The abundance of Ruminococcaceae family, Coprococcus_1 genus and Blautia genus were significantly decreased with psoriasis improvement, which had been demonstrated significantly increased in psoriasis. (3) Mice receiving psoriatic microflora transplantation showed significantly delayed recovery of psoriasiform dermatitis and less reduction of IL-17A, than those receiving healthy microflora or blank control (p<0.05 and p<0.01, respectively).

**Conclusions:** Multiple evidences we provided here demonstrate the involvement of gut microbiota in psoriasis development. The strategy based on gut microbiota is expected to be a promising supplementary for long-term management of psoriasis.

## 1. Introduction

Psoriasis is a common chronic skin inflammation, and it can even cause systemic involvement for those with early-onset and severe conditions.^1^ Although the exact pathogenesis is not completely known, psoriasis has been considered a relapsing-remitting disease triggered by environment-immunity interaction in genetically susceptible individuals.

Treatment options have advanced following deeper understanding of the pathophysiology of psoriasis, e.g. IL-23/IL-17-targeted agents. However, a survey from the National Psoriasis Foundation reveals widespread treatment dissatisfaction in psoriatic patients (52.3%),^2^ especially in reducing the recurrence and managing its long-term chronic course.

The gut microbiota, 100 trillion microorganisms residing in the human gastrointestinal tract, has been documented to provide essential benefits to host health, particularly by orchestrating immune/inflammation homeostasis.^3^ Evidence suggests that lower gut microbiome diversity is associated with higher levels of fat and low-grade chronic inflammatory process.^4^ Dysbiosis in gut microbiota has been implicated in continuous immunological stimulation, as a trigger for local and (or) systemic immune responses, including in inflammatory bowel disease (IBD) and allergy.^5,6^

Accumulating evidence has suggested the association between dysbiosis of gut microbiota and psoriasis. (1) The epidemiological association between psoriasis and IBD showed increased prevalence of IBD in psoriatic patients, and *vice versa*.^7,8^ (2) The partial shared susceptibility loci and DNA polymorphisms between psoriasis and IBD (e.g. 6p21.3) further supports their association at genetic level.^9,10^ (3) Notably, psoriatic patients have been shown decreased bacterial diversity and changed relative abundance of certain bacterial taxa, resembling dysbiosis in IBD.^11-15^ (4) The microbiota profile in severe psoriasis has been demonstrated different from those with mild one.^16^ By now, elucidating gut microbiota status and the cross-talk of microbiota & immune system in psoriatic patients are at their initial stages. It will provide theoretical basis to develop promising microbiome-based therapeutic options.

The purpose of current study is to further strengthen the involvement of gut microbiota in psoriasis development by epidemiological investigation of gastrointestinal discomfort symptoms in psoriatic patients, and by the analysis of gut microbiota variation with psoriatic improvement. And we also analyzed the recovery status and pathogenic cytokines (e.g. IL-17A) in mouse psoriasiform models that were transplanted with fecal microbiota from psoriatic patients or healthy controls. Multiple evidences we provided here demonstrated the involvement of gut microbiota in psoriasis development. The strategy by manipulating gut microbes is expected to be a promising supplementary therapeutic method for the long-term management of psoriasis.

## 2. Materials and Methods

### 2.1. Ethical statement

All human experiments and animal experiments were approved by the Ethics Committee of Sichuan Provincial People’s Hospital. Written informed consent of all psoriatic patients and healthy controls were obtained.

### 2.2. Epidemiological investigation

This epidemiological survey was performed from January 2018 to June 2020 to investigate the prevalence and severity of gastrointestinal discomfort symptoms in psoriatic patients and general population by the questionnaire (Appendix S1). The details of the inclusion and exclusion criteria are in Table S1.

### 2.3. Patients and fecal samples

#### 2.3.1 Psoriatic patients (un)treated with Acitretin and their fecal sample processing

Patients with moderate to severe vulgaris psoriasis from outpatient clinic of the department of Dermatology were included. The general inclusion and exclusion criteria were showed in patients section of Table S1. Patients in group with Acitretin treatment had been orally administered with Acitretin Capsules for one month at a dose of 0.5 mg/kg/d (Huapont Pharmaceutical Co., Ltd, Chongqing, China). Patients improved more than 75% were enrolled randomly, based on their PASI (Psoriasis area and severity index) score. Untreated psoriatic patients with matched age and gender served as controls. Fecal samples of all patients were collected in the morning, and immediately stored at −80°C for 16S rRNA analysis. The whole collection procedure was completed within 30 minutes.

#### 2.3.2 Participants and their fecal sample processing for fecal microbial transplantation (FMT)

Fecal samples were collected from four patients with moderate to severe psoriasis (2 males and 2 females) and four age- and gender-matched non-psoriatic controls. All these voluntary participants were 18-45 years old, and other requirements are in accordance with Table S1. About 20g of feces were freshly collected from each participant in the morning. They were divided into five aliquots after removal of undigested solids within 30 min of collection. Each 0.5g was stored in a sterile storage tube at −80°C.

#### 2.3.3 Processing mouse fecal sample for 16S rDNA sequencing analysis

All animal experiments were conducted in accordance with National Institutes of guidelines for animal care and use. In order to confirm the successful FMT, characteristics of mouse gut microbes before and after FMT were analyzed by 16S rDNA sequencing technology. Following slightly pushing mouse lower abdomen using a moist cotton swab to provoke defecation in the morning, a minimum of five fresh fecal pellets were collected in sterile storage tubes and immediately kept at −80°C. The feces were collected before FMT (pre-FMT), at 0 day and 4th day after complete FMT procedure respectively.

### 2.4 DNA extraction and 16S rRNA amplification sequencing analysis

Metagenomic DNA was isolated from human and mouse samples using CTAB methods and QIAamp 96 PowerFecal QIAcube HT kit (QIAGEN, Germany) following the manufacturer’s instructions respectively. The amplifications of V4 (human samples) and V3&V4 (mouse samples) regions of bacterial 16S rRNA gene were performed by PCR using the barcoded primers of 515F&806R (human samples) and 343F&798R (mouse samples) respectively. Amplicons were further purified with GeneJET Gel Extraction Kit (Thermo Scientific, USA) and pooled together. All purified samples were sequenced on the Illumina Miseq platform (Illumina Inc., CA) with generating 300 bp paired-end reads.

### 2.5 Microbial profiling analysis

All raw sequencing data were in FASTQ format. Trimmomatic software was used to trim raw sequence that cutting off ambiguous bases and base quality below 20 found after sliding window trimming approach.^17^ Contiguous sequences were then assembled by FLASH software.^18^ Operational taxonomic unit (OTU) tables with 97% nucleotide identity were constructed under the condition that sequences were performed further denoising using QIIME software (version 1·8·0).^19^ The representative read of each OTU were annotated and blasted against Greengenes database.^20^

The Shannon’s diversity, Simpson diversity index, Chao1 index and Abundance-based Coverage Estimator (ACE) were calculated to estimate the within-community diversity and richness of the gut microbiota. Based on alpha diversity metrics, rarefaction curves were generated and drawn by “vegan” package in R (Version 2·15·3) to assess depth of coverage. UniFrac distances between bacterial communities were calculated on a phylogenetic tree, and unweighted results were represented in Principal Component Analyses (PCoA) or Nonmetric Multidimensional Scaling (NMDS) using R software (Version 2·15·3).^21^ And results of Euclidean distance were depicted in NMDS. Differentially abundant taxa between groups was identified by MetaStat and Linear discriminant analysis (LDA) coupled with logarithmic LDA score cutoff of 4·0.^22^ Metastats analysis was performed by using R software (Version 2·15·3), P<0.05 was set as significant threshold.

### 2.6 Imiquimod-induced psoriasiform dermatitis model in mice

The 8-week-old female C57BL/6 mice (18-20g, Chengdu Dossy Experimental Animals, Sichuan, China, certification No. SCXK chuan 2015-030) were fed with free access to food and water under specific pathogen-free conditions. A 2cm × 4cm area of dorsal skin of mice was shaved and depilated. A daily topical dose of 62.5 mg of listed imiquimod (IMQ) cream (5%, Mingxinlidi Laboratory, China) was then applied on the hair-free back for five consecutive days to induce psoriasiform dermatitis. The severity of skin inflammation was evaluated by scores of the skin scaling and erythema (0 to 4 respectively).^23^

### 2.7 Transplanting human fecal microbiota into psoriasiform dermatitis model

After thawed for about 15 minutes on ice, 2g frozen fecal samples from patient group or healthy group (0.5g from each donor) were pooled and diluted in sterile reduced phosphste-buffered saline (PBS, 0.1M, PH 7.2) at 200 mg/ml. After vortex, the suspension was passed through 0.5 mm stainless steel laboratory sieves to remove large particulates and fibrous matters. Fecal suspension (200 μl) from psoriatic patients (P group) or healthy controls (N group), or blank control (PBS, C group) were given by oral gavage into psoriasiform dermatitis mouse model once per day for consecutive three days.

### 2.8 Tissue collection and H&E staining

Mice were sacrificed by cervical dislocation under anesthesia with 1% sodium pentobarbital solution on Pre-FMT, Day 0 after FMT, and Day 4 after FMT. The 0.5cm×0.5cm skin lesions were gently removed and rinsed with physiological saline. They were immediately formalin-fixed (4%) and embedded in paraffin. Hematoxylin and eosin (H&E) staining was performed routinely. Epidermal thickness was evaluated under three high-power fields of light microscope (NIKON ECLIPSE CI, Japan) by three independent researchers.

### 2.9 Immunofluorescence studies

After routine processing and blocking, sections were incubated at 4°C overnight with anti -mouse primary antibodies against TNF-α (RRID:AB_2835319), IFN-γ (RRID: AB_10857066), IL-17A (RRID: AB_2838094), IL-17F (RRID:AB_2842177), IL-23(RRID: AB_10852886) and FOXP3 (RRID: AB_2861434) or isotype control respectively. After rinsing, sections were treated with Cy3-conjugated goat anti-rabbit IgG secondary antibody (RRID: AB_2861435) for two hours, and then counterstained with DAPI. Image acquisition was performed with a digital slide scanner (3DHISTECH, Budapest, Hungary) under ECLIPSE TI-SR fluorescent microscope (NIKON, Japan). Positive immune cells and their values were determined to assess inflammatory changes.

### 2.10 Statistical analysis

Numerical results are expressed as median with a 95% confidence interval. Categoric variables were described with numbers and percentages. Differences of BMI and age between two groups were compared with Mann-Whitney U Test. The proportions among patients and controls were compared by chi-squared test. The relationship between gastrointestinal symptoms and psoriasis was evaluated by a logistic regression test. P<0.05 was considered as a statistically significant difference.

## 3. Results

### 3.1. Higher incidence of gastrointestinal discomfort symptoms in psoriatic patients than common population

Totally 459 participants returned their questionnaires, and 326 were qualified, including 167 psoriatic patients (115 males and 52 females) and 167 non-psoriatic controls (84 males and 83 females). The two groups were age-matched, and the differences in sex ratio and Body Mass Index (BMI) were consistent with previous epidemiological findings. The summary of demographic and clinical details was described in Table 1.

**Table 1.**
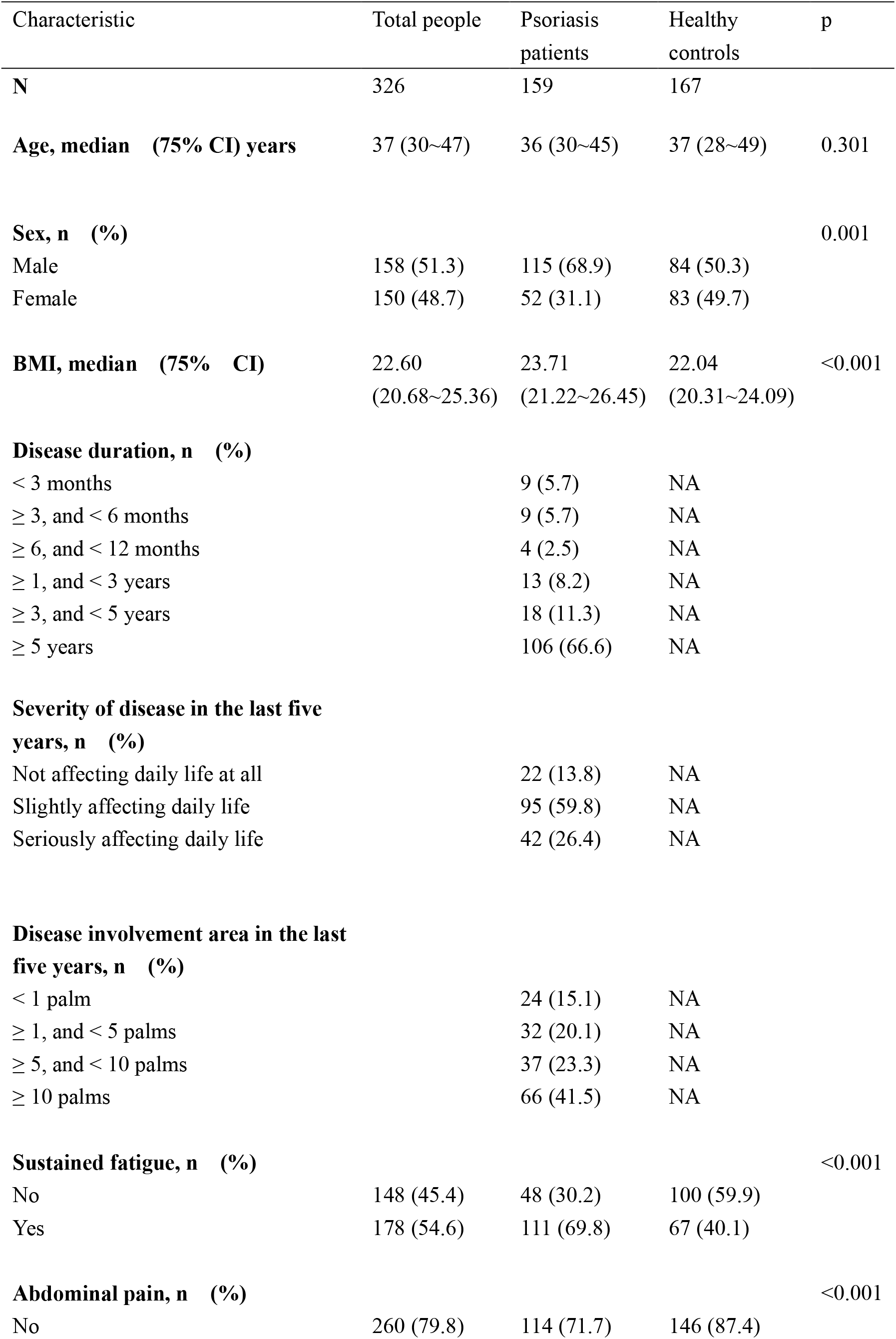
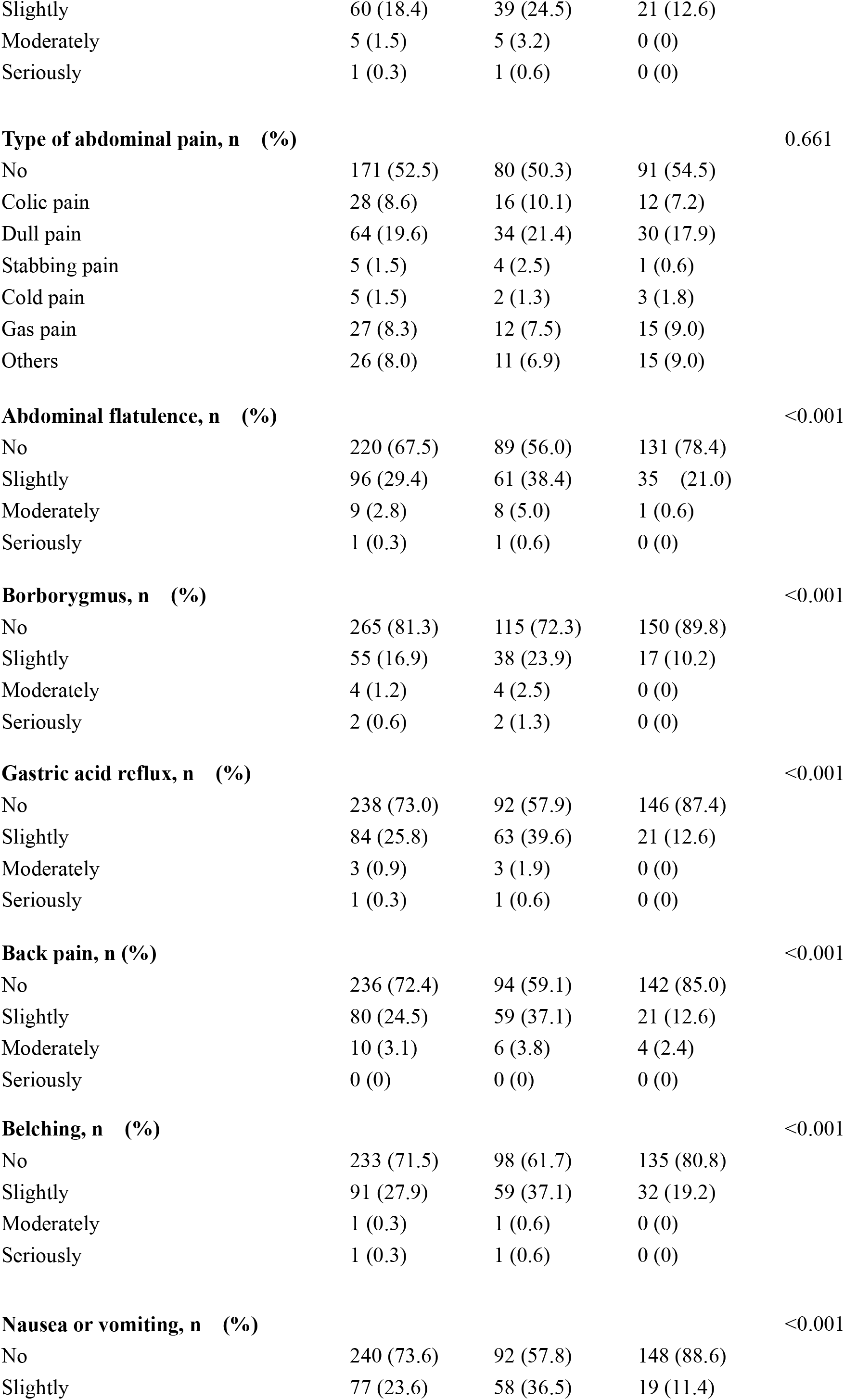
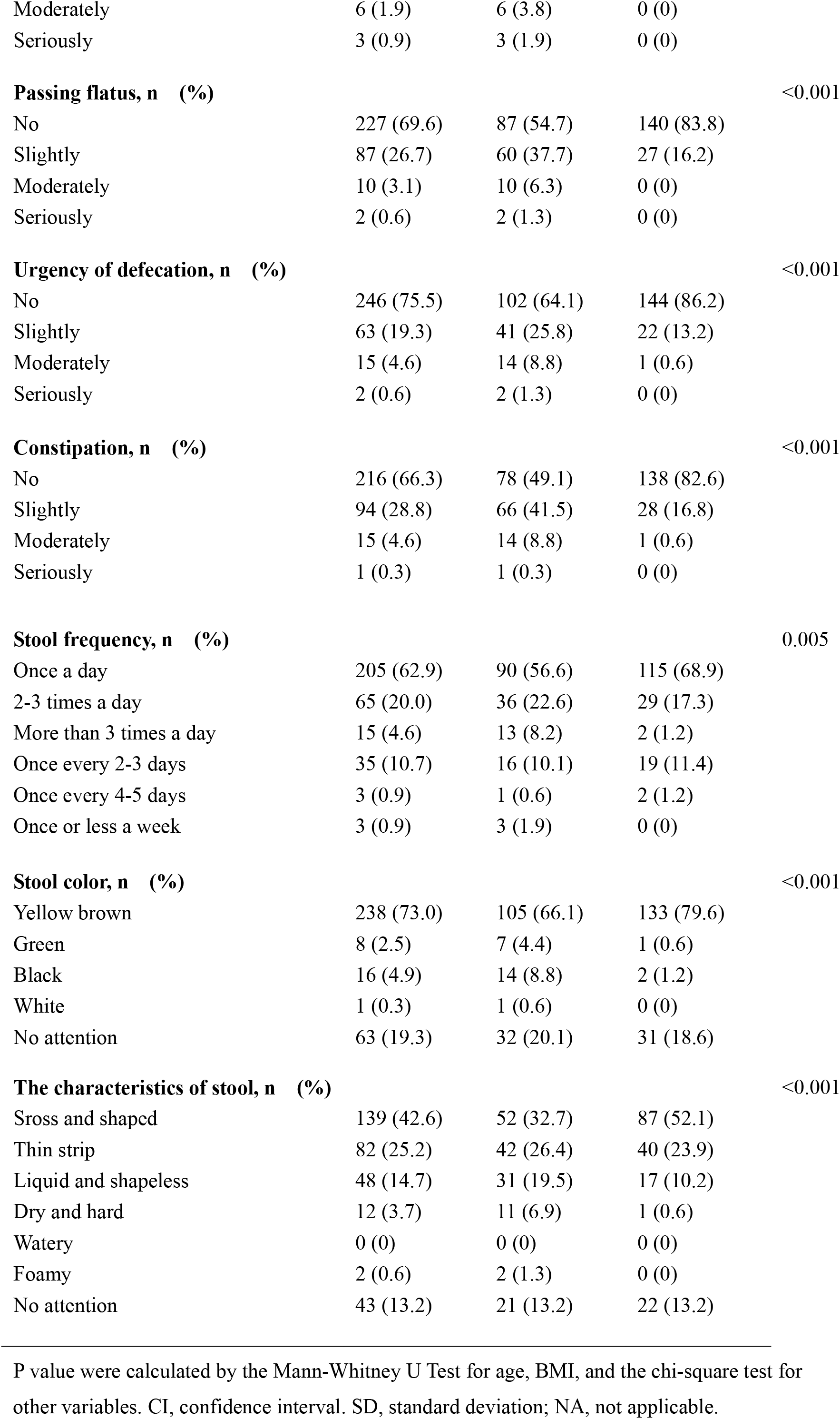
Demographic information and gastrointestinal discomfort symptoms in psoriatic patients and common population.

According to this investigation, 85.53% of psoriatic patients *versus* 58.08% of common controls presented with at least one gastrointestinal symptom. The prevalence of the symptoms, including sustained fatigue, abdominal pain, abdominal flatulence, borborygmus, gastric acid reflux, belching, nausea or vomiting, passing flatus, urgency of defecation, and constipation, was significantly higher in psoriatic patients, compared with common population (p<0.01, Table 1). There were significantly different proportion of patients and controls with abnormal stool frequency (20.75% *vs* 13.77%), stool color (13.84% *vs* 1.80%), and characteristics of stool (54.09% *vs* 34.73%).

Results of logistic regression analysis showed that sustained fatigue, passing flatus, and constipation were significantly correlated with psoriasis (p<0.05, Table 2). Patients with psoriasis were more likely to experience these discomfort symptoms, which suggested possible involvement of gastrointestinal tract in psoriasis development.

**Table 2.** Results of Logistic regression analysis of gastrointestinal symptoms in psoriasis.

| **Variables** | **Odds ratio** | **p** |
| --- | --- | --- |
| **Age** | 0.97 (0.94~1.00) | 0.066 |
| **Sex** | 1.41 (0.75~2.66) | 0.287 |
| **BMI** | 1.13 (1.03~1.23) | 0.007 |
| **Sustained fatigue** | 1.96 (1.05~3.63) | 0.034 |
| **Abdominal pain** |  | 0.504 |
| No | 0.49 (0.20~1.22) |  |
| Slightly | NA |  |
| Moderately | NA |  |
| **Abdominal distension** |  | 0.757 |
| No | 1.30 (0.63~ 2.72) |  |
| Slightly | 0.84 (0.06~ 11.99) |  |
| **Borborygmus** |  | 0.964 |
| No | 1.28 (0.52~ 3.16) |  |
| Slightly | NA |  |
| Moderately | NA |  |
| **Gastric acid reflux** |  | 0.912 |
| No | 1.35 (0.60~ 3.06) |  |
| Slightly | NA |  |
| Moderately | NA |  |
| **Backache** |  | 0.556 |
| No | 1.53 (0.69~ 3.38) |  |
| Slightly | 0.82 (0.10~ 6.94) |  |
| **Belching** | 1.00 (0.51~1.95) | 0.990 |
| **Nausea and vomiting** | 1.55 (0.75~3.21) | 0.242 |
| **Increased fart** |  | 0.023 |
| No | 2.95 (1.49~5.84) |  |
| Slightly | NA |  |
| Moderately | NA |  |
| **Diarrhea** |  | 0.668 |
| No | 0.66 (0.26~ 1.67) |  |
| Slightly | 2.58 (0.22~ 30.62) |  |
| Moderately | NA |  |
| **Constipation** |  | 0.021 |
| No | 3.04 (1.40~ 6.61) |  |
| Slightly | 19.67 (0.10~ 389.35) |  |
| Moderately | NA |  |
| **Defecation frequency** |  | 0.445 |
| Once a day | 0.91 (0.43~ 1.93) |  |
| 2-3 times a day | 4.02 (0.49~ 32.91) |  |
| More than 3 times a day | 0.41 (0.13~ 1.28) |  |
| Once every 2-3 days | 0.19 (0.01~ 4.58) |  |
| Once every 4-5days | NA |  |
| **Fecal color** |  | 0.174 |
| Yellow brown | 8.60 (0.66~ 112.14) |  |
| No attention | 4.22 (0.68~ 26.14) |  |
| Green | NA |  |
| Black | 1.93 (0.86~ 4.32) |  |
| **Fecal traits** |  | 0.929 |
| Sross and shaped | 0.90 (0.43~1.90) |  |
| Thin strip | 1.00 (0.38~ 2.61) |  |
| Liquid and shapeless | 4.02 (0.32~ 50.50) |  |
| Dry and hard | NA |  |
| No attention | 1.06 (0.40~ 2.81) |  |

NA, not applicable, due to the insufficient number of cases available for the statistics.

### 3.2 Variation of gut microbiota accompanied with psoriasis improvement

Gut microbiota in psoriatic patients and healthy controls has been already demonstrated to be significantly different.^16,24^ To further understand the relationship between gut microbiota and psoriasis, we then investigate the variation in bacteria community of psoriatic patients who were effectively treated by Acitretin.

A total of 20 patients (10 untreated and 10 treated with Acitretin) of moderate to severe vulgaris psoriasis were included (Table S2). The gastrointestinal bacterial diversity and composition was evaluated by pyrosequencing analysis based on 16s rRNA. The coverage of applied sequencing depth was adequate, as indicated by goods coverage rarefaction curves of two groups which tend to be plateau (Fig S1A). The results of Alpha diversity indexes indicated similar community richness and species diversity in both groups (p=0.545 for Chao; p=0.112 for Simpson, Fig S1B and S1C). We further applied NMDS to assess the differences of microbial communities between two groups and found that most of the Untreated group were discriminated from the majority of all samples (Fig. 1A), although adonis analyses revealed no significant differences between two groups (P=0.382).

**Figure 1.**
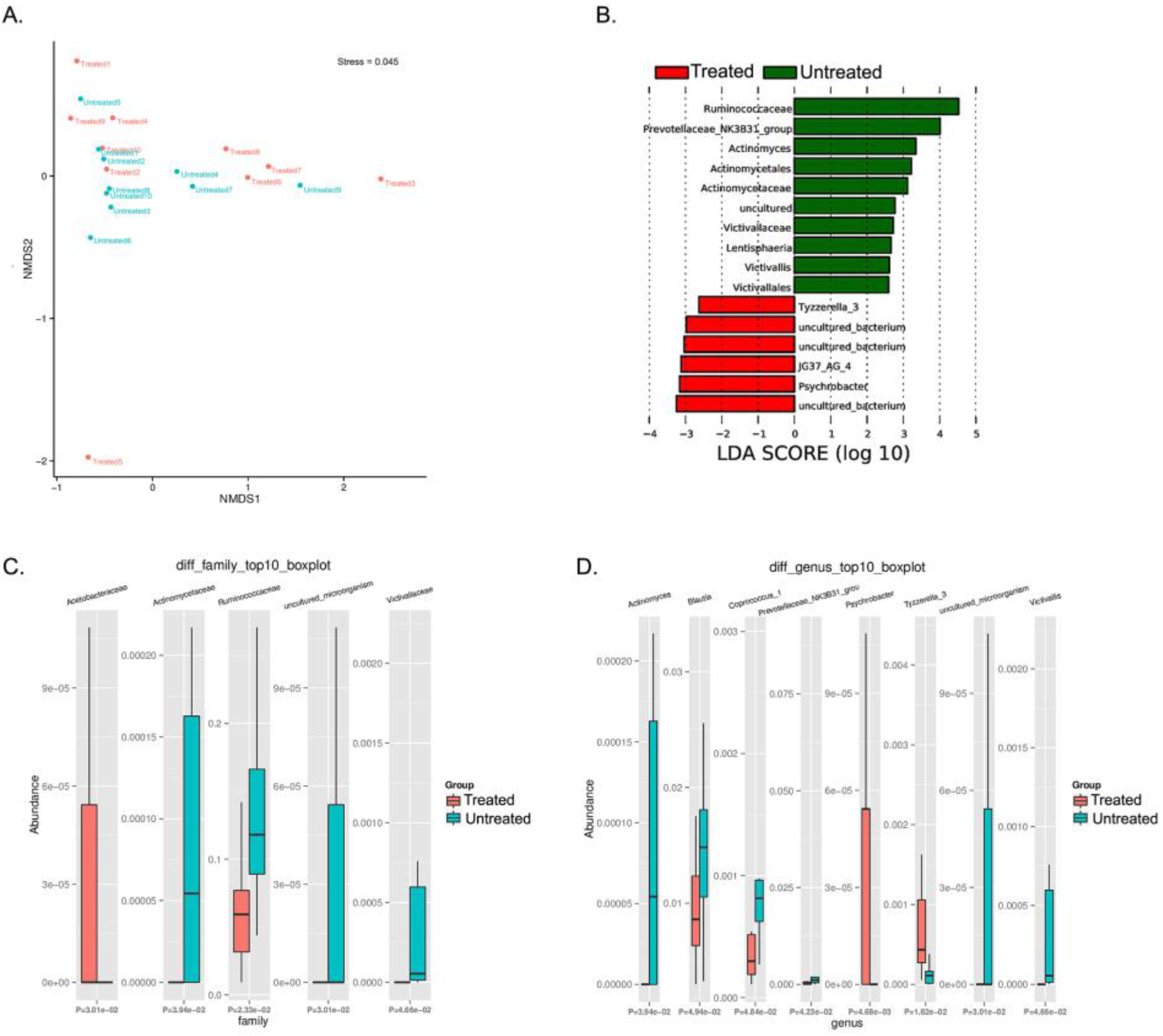
Taxonomic composition of bacterial community in psoriatic patients (un)treated with Acitretin. (A) Nonmetric Multidimensional Scaling (NMDS) analysis with unweighted UniFrac displayed that most of the Untreated group were discriminated from the majority of Treated group samples. Each colored solid circle represents one sample. Solid circles that are closer together represent similar taxonomic composition. (B) The scores of linear discriminant analysis for the differentially abundant taxa. Significant bacterial differences at family level (C) and at genus level (D) between two groups were analyzed by Metastats.

Taxonomic composition revealed differences in the abundance of specific bacterial cluster by LefSe analysis and kruskal-wallis analysis (Fig. 1B, 1C, and 1D). *Bacteroidetes, Firmicutes* and *Proteobacteria* phylum were dominant in two groups with similar abundance (Table S3). Within identified bacterial components of other taxonomic levels, the comparison between the two groups rendered a significantly increased abundance of *JG37_AG_4* class, *Acetobacteraceae* family, *Psychrobacter* genus and *Tyzzerella_3* genus in Treated group. Moreover, there were more bacteria with significantly decreased relative abundance in Treated group, including *Lentisphaeria* class, as well as *Victivallales* and *Actinomycetales* at the order level, and *Ruminococcaceae, Actinomycetaceae* and *Victivallaceae* at the family level. At the genus level, *Actinomyces, Prevotellaceae_NK3B31_group, Victivallis, Coprococcus_1* and *Blautia* were also significantly decreased in Treated group.

Published studies comparing intestinal bacteria in psoriatic patients and healthy controls have indicated specific bacterial differences at all levels of taxonomic classification (Table 3).^11,13,14,16,24-27^ The variations in certain bacteria presented in our study here are consistent with the results of these previous studies. Specifically, significantly decreased *Ruminococcaceae* family, *Coprococcus_1* genus and *Blautia* genus in Treated group corresponds to their significantly increased relative abundance in psoriasis group compared with healthy people. Of them, *Blautia* genus is related to anti-inflammatory properties.^28^ In brief, the relative abundance of certain “psoriatic characteristic microbiota” was reduced in patients after treatment, suggesting that gut microbiota was closely associated with psoriasis development.

**Table 3.**
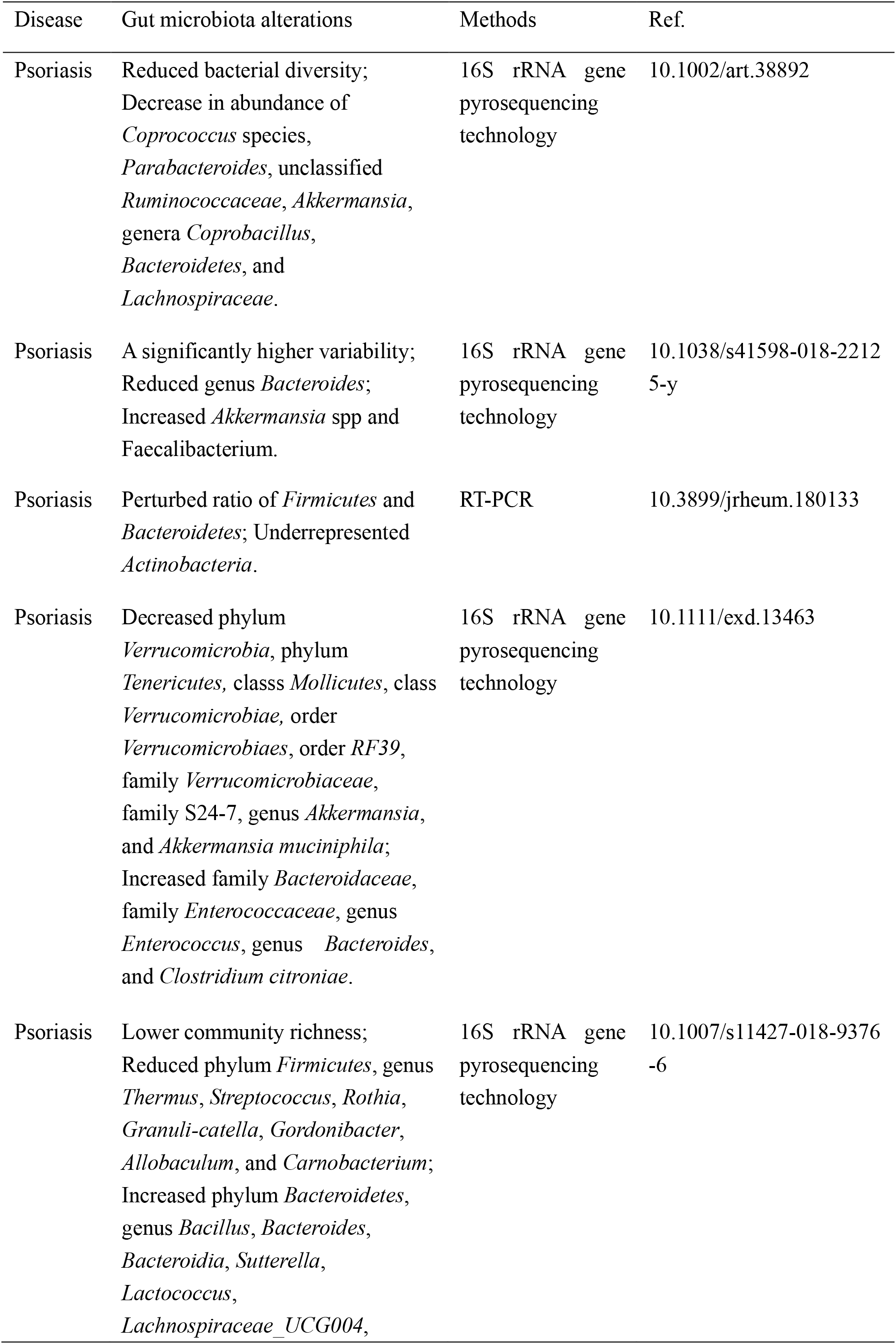
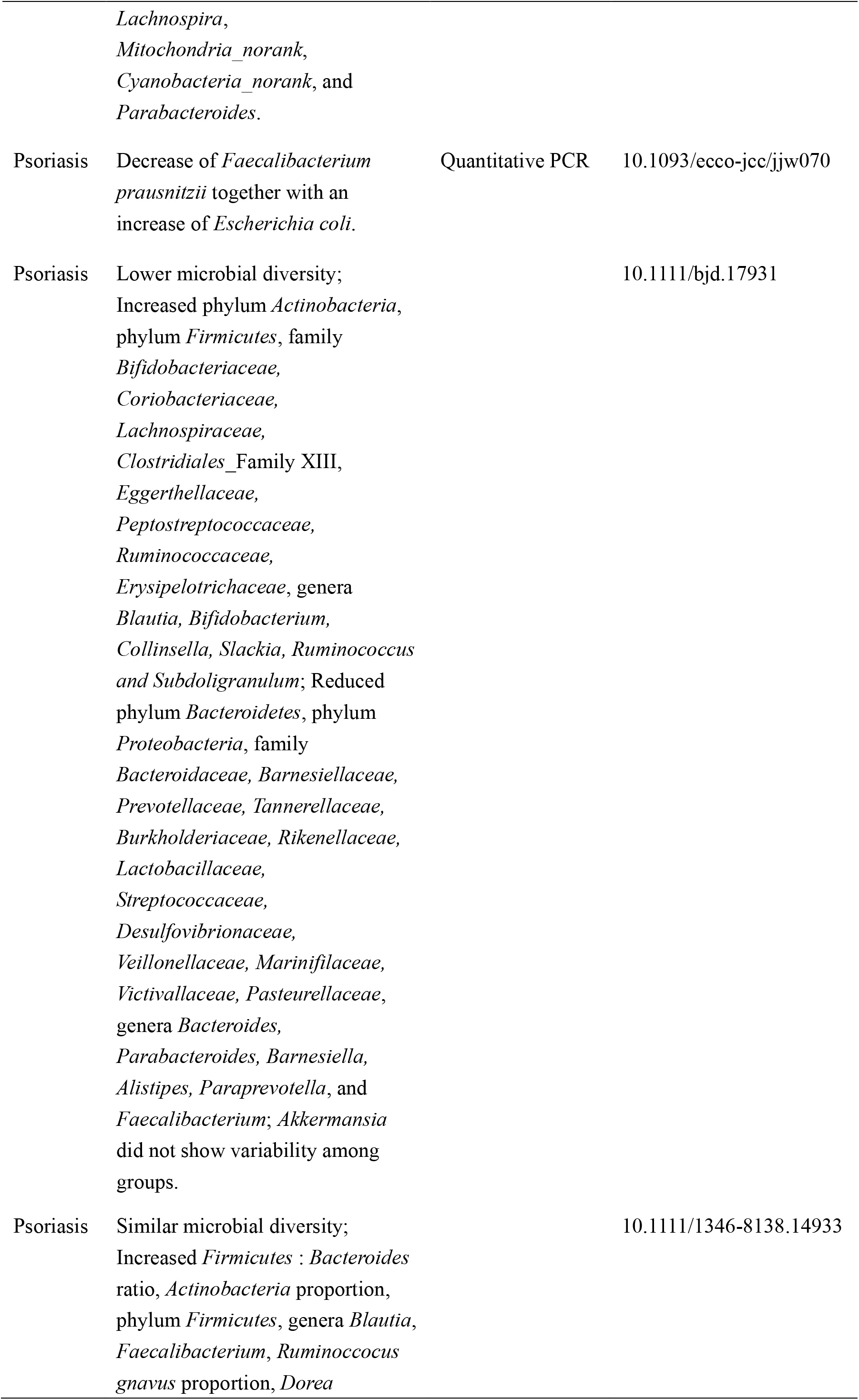
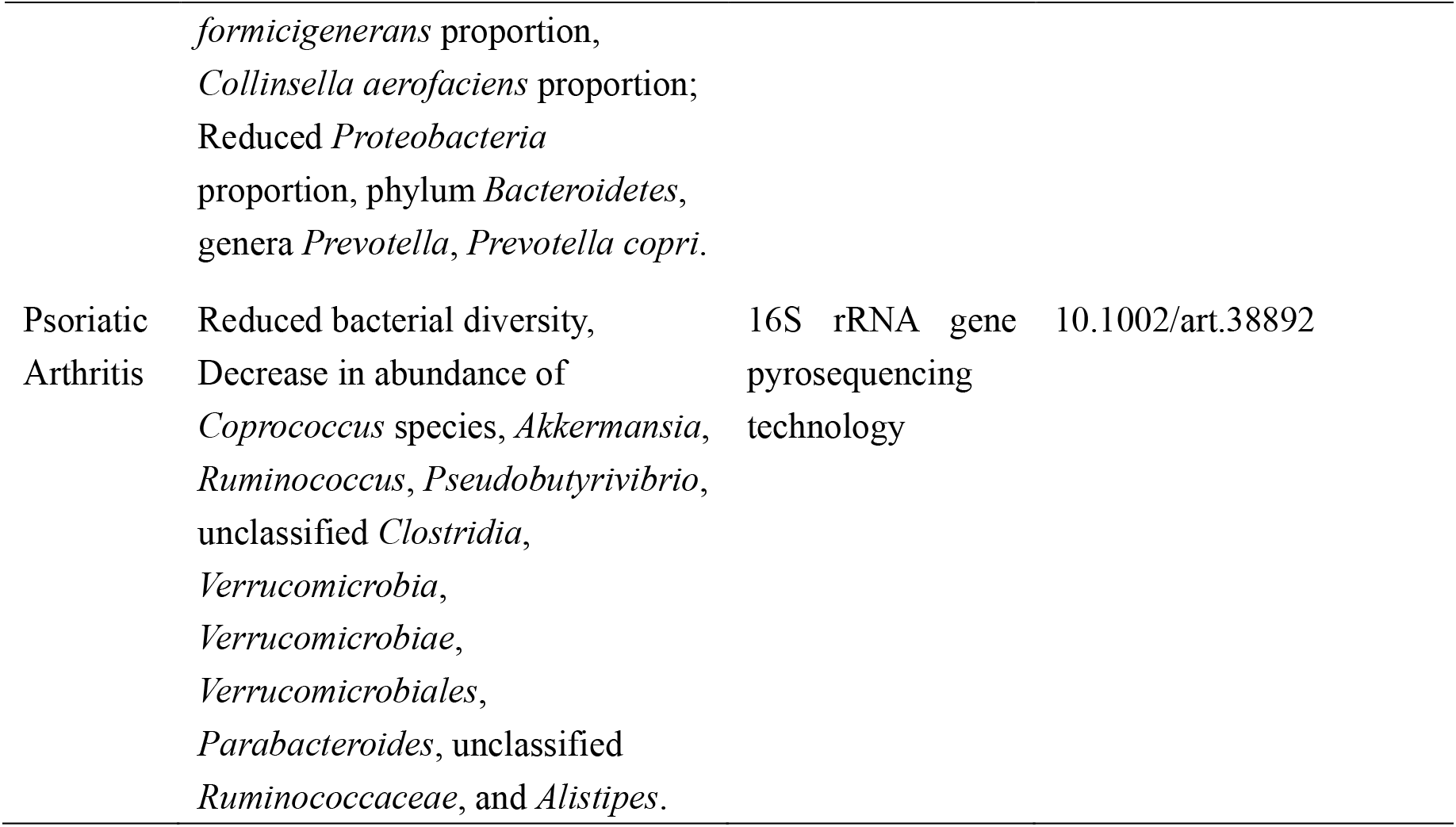
Gut microbial alterations in patients with psoriasis/psoriatic arthritis compared with normal controls, based on published literatures.

### 3.3 Significantly delayed recovery of psoriasiform dermatitis in mice receiving psoriatic microflora transplantation

We first confirmed the successful transplantation of human fecal microbiota into mouse psoriasiform models by analyzing the inner structure of mouse microbial community at different time-point (pre-FMT, at 0 and 4^th^ day after complete FMT procedure). Chao1 and Shannon indexes of all samples were calculated (Table S4). Although pre-FMT group and each group at day 0 after FMT displayed similar Chao1index (Fig. S2A, 2B, and 2C), microbial diversity in PFM-0d group or NFM-0d group was significantly decreased respectively, compared with Pre-FMT group, as determined by Shannon index (p<0.01, Fig S2D, S2E and S2F). The results of PCoA based on unweighted UniFrac distance revealed significant clustering of seven groups from each other (Supplementary Fig S2G). Notably, adonis analyses further revealed that overall microbiota structure significantly differed among seven groups (p<0.05, Table S5). These data confirmed that human fecal microbiota were transplanted into mouse psoriasiform models successfully.

IMQ cream applied onto the shaved back skin of mice can induce skin inflammation accompanied by human psoriasis-like pathological features, including remarkable acanthosis, parakeratosis and infiltration of inflammatory cells in the superficial dermis. After termination of one-week IMQ application, typical psoriasis-like phenotype could relief or even disappear over time. Here we investigated the effects of different fecal microbial transplantation (from psoriasis patients or healthy people) on the course of IMQ-induced mouse psoriasiform dermatitis.

Typical lesions with erythema, scaling and thickening were observed after one-week IMQ application, compared with normal mouse skin (Fig 2A, pre-FMT). The cumulative score (erythema plus scaling) was depicted in Fig 2B. At Day 0 after three consecutive days of FMT, PFM-0d group had significantly higher scores than NFM-0d group or CON-0d group (p<0.05), and their higher severity of psoriasis-like clinical characteristics was observed. The psoriatic manifestations in all groups were almost disappear at Day 4 after FMT, when PFM-4d group, NFM-4d group and CON-4d group showed similar severity.

**Figure 2.**
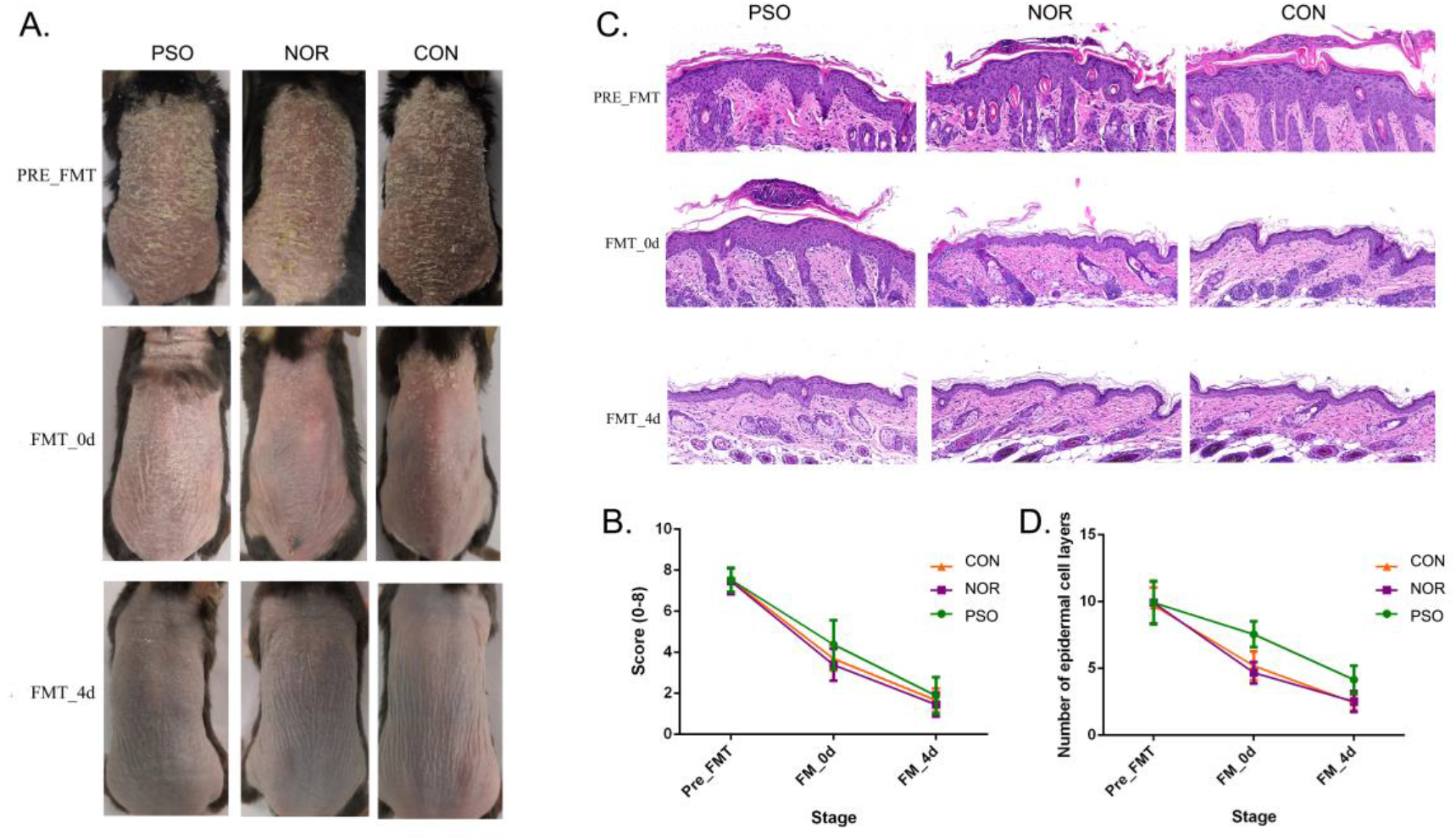
Mice received FMT from psoriatic patients showed significantly delayed recovery of psoriasiform dermatitis. After daily application of IMQ cream for five consecutive days, mice in the different groups were respectively transplanted with fecal microbiota from psoriatic patients (PSO), healthy controls (NOR), or control of PBS (CON). (A) Phenotypic presentation of dorsal skin of mice in different group. (B) Scores of skin lesions were calculated by erythema plus scaling (a scale from zero to four, respectively). (C) H&E staining (×100) of dorsal skin of mice from different group. (D) Epidermal thickness was indicated by number of epidermal cell layers. Colored symbols in C and D indicated mean score ± SD of five mice per group.

Analysis of corresponding pathological slices from IMQ-induced psoriasiform dermatitis revealed significant increases in epidermal thickening in PFM-0d group, compared with NFM-0d group or CON-0d group (p<0.01, Fig 2C, 2D). Differences in epidermal thickening among three groups at Day 4 after FMT were highly comparable to that at Day 0 after FMT. Although there are no significant differences between the two groups, the NFM group presented slightly reduced thickness of the epidermis, compared with the CON group. Thus, mice receiving FMT from psoriatic patients displayed more delayed recovery of psoriasis-like phenotype up to the end of the experiment. These results showed a close association between gut microbiota and distal skin inflammation, indicating that changes in gut microbiota may have a significant impact on the course of psoriasis.

### 3.4 Less reduction of IL-17A in mice transplanted with psoriatic fecal microbiota

To further verify the possible mechanism of gut microbiota on distal skin changes, we analyzed expression levels of cytokines, including IL-17A, IL-17F, IFN-γ, TNF-α, IL-23 and transcription factor Foxp3 in psoriasiform mice. The immunofluorescence results showed that IL-17A expression in mouse skin gradually decreased after the termination of IMQ application, accompanied by gradually improved psoriasiform skin lesions (Fig 3). The level of IL-17A in PFM-0d group was significantly higher than that in NFM-0d group or CON-0d group, suggesting its less reduction (p<0.01). Although the expression of IL-17A did not differ significantly among three groups at Day 4 after FMT, its expression was relatively high in PFM-4d group.

**Figure 3.**
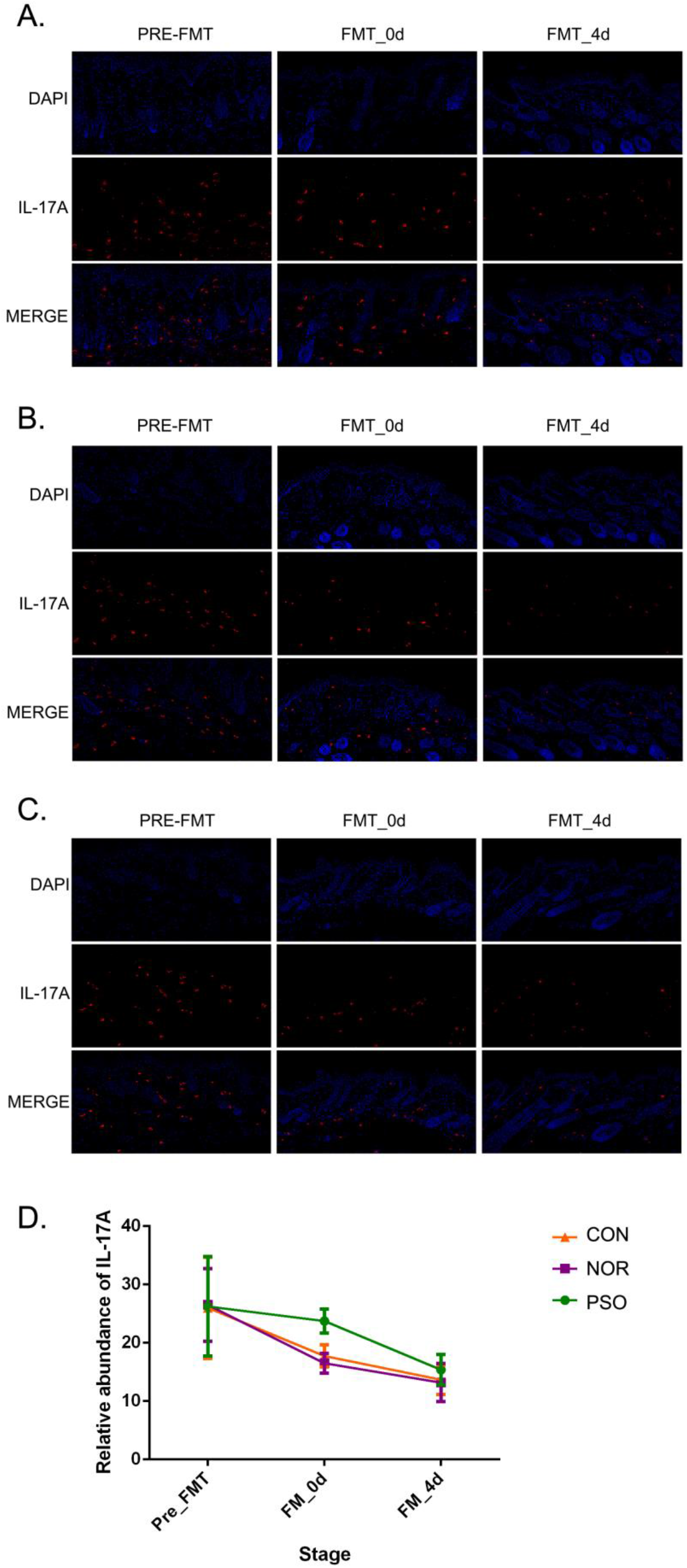
Analysis of IL-17A in mouse skin lesions of psoriasiform by immunofluorescence assay. (A) mice received FMT of psoriatic fecal sample. (B) mice received FMT of normal fecal sample. (C) control mice received oral gavage of PBS. Blue fluorescence represents DAPI; Red fluorescence represents IL-17A. (D) Numbers of red fluorescence were counted to analyze IL-17A expression. Colored symbols indicate mean number ± SD of five mice per group.

In order to explore the mechanism underlying the less reduction of IL-17A in lesions and considering the possible role of gut-skin axis, we analyzed the changes of IL-17A in gastrointestinal tissues. There was relatively low level of IL-17A expression in gastrointestinal tissues of psoriasiform mice before FMT, and we showed that FMT had an effect on its level (Fig 4). In N and C group, IL-17A increased slightly at Day 0 after FMT, and its expression at Day 4 after FMT was almost the same as that at pre-FMT (Fig 4D). No significant differences between N and C group was observed in all time-points (p>0.05). It should be noted that IL-17A expression in P group was significantly increased after FMT (p<0.01), compared with that in N or C group (Fig 4). This increase corresponds to the less reduction of IL-17A in lesions. There was no such a corresponding relationship for other cytokines of IL-17F, IL-23, IFN-γ, TNF-α and Foxp3 transcription factor (data not shown).

**Figure 4.**
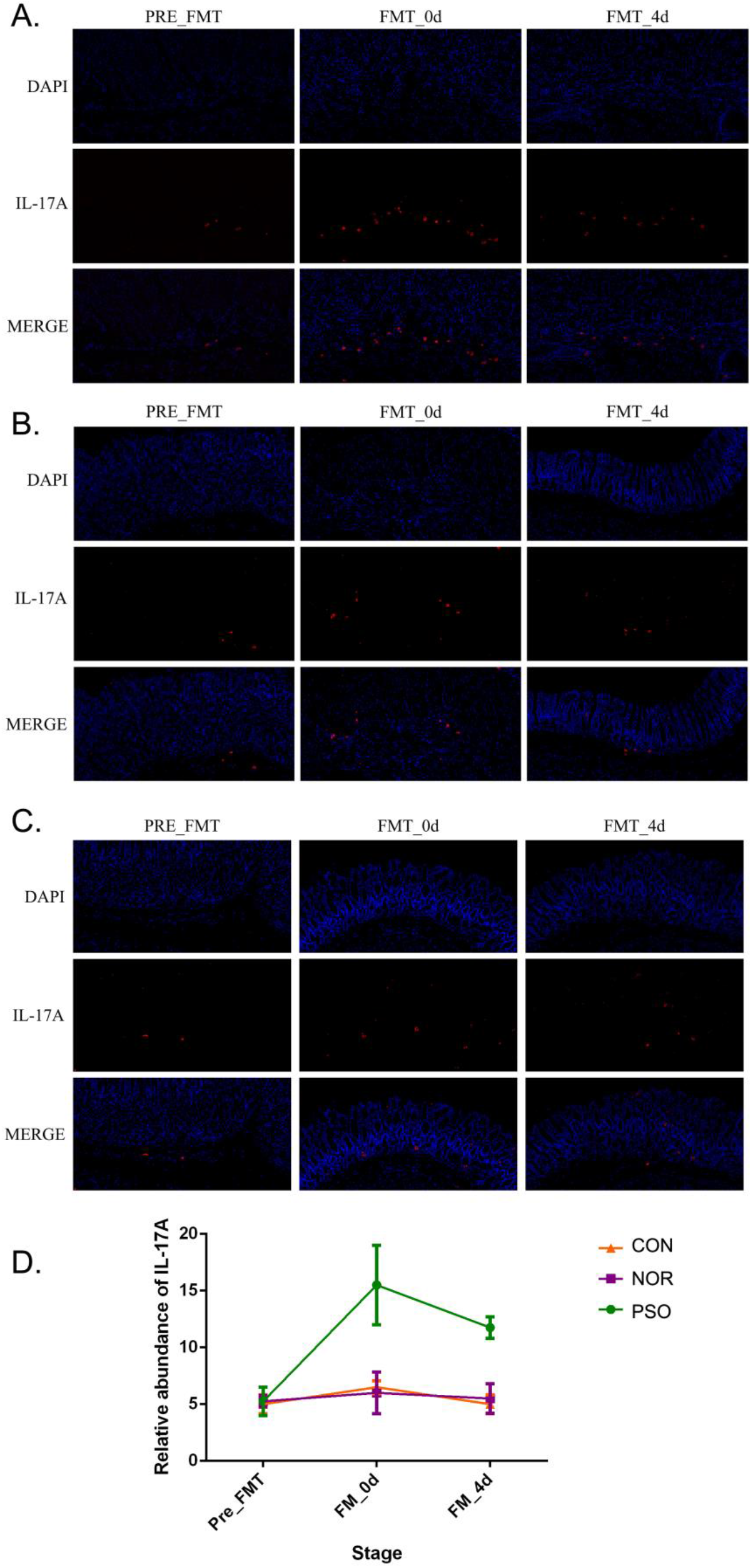
Analysis of IL-17A in mouse gastrointestinal tissues of psoriasiform models by immunofluorescence assay. (A) mice received FMT of psoriatic fecal sample. (B) mice received FMT of normal fecal sample. (C) control mice received oral gavage of PBS. Blue fluorescence represents DAPI; Red fluorescence represents IL-17A. (D) Numbers of red fluorescence were counted to analyze IL-17A expression. Colored symbols indicate mean number ± SD of five mice per group.

## 4. Discussion

Evidences associating gut bacteria with distant extra-intestinal inflammation (e.g. the skin) through regulation of immune system have been expanding.^29-31^ The clinical observations on psoriatic comorbidities (e.g. IBD) and alterations in architecture of intestinal barrier have fueled the study concerning the correlation between psoriasis pathogenesis/development and gut microbiota.

The present study demonstrated the involvement of gut microbiota in the course of psoriasis from several aspects. We first found that multiple gastrointestinal symptoms were significantly more frequent in psoriatic patients than common controls. Except for gastric acid reflux, other gastrointestinal symptoms involved in this study were not found to be related to gender. Therefore, gender imbalance has little effect on the difference in gastrointestinal symptoms between the two groups. Next, we found the recovery process of psoriatic patients was accompanied by significant reduction of certain “psoriatic characteristic microbiota”,^11,13,14,25^ which had been identified by comparing gut bacteria in psoriatic patients and healthy controls. This alteration of characteristic microbiota signature might be attributed to Acitretin itself or be related with the recovery of psoriasis directly. We last showed significantly delayed recovery of psoriasiform dermatitis in mice receiving psoriatic microflora transplantation, compared with those receiving healthy microflora. These investigations, observations, and previous published data suggest manipulation of gut microbiota, such as healthy microflora transplantation, could be a supplementary option for psoriasis treatments. This manipulation should aim to targeting the whole community rather than focusing on certain taxa, considering the complex interactions among bacterial microorganisms.

Recipient mice used in this study were not germ-free or treated with antibiotics like previous literatures reported. Although avoiding interference of their own intestinal microbiota, germ-free and antibiotics-treated mice have some limitations. Antibiotics could not only affect systemic immunity, but also limit the colonization of donor microbiota. Similarly, germ-free mice with abnormal intestinal structure have been reported to secrete more lipids resulting in more susceptible to low-grade inflammation or even imbalanced cytokines and immune cells.^32^

In terms of mechanism, there was significantly increased IL-17A expression in the gastrointestinal tract of mice receiving psoriatic microflora transplantation, which correspondingly showed significantly delayed deduction of IL-17A in lesions and significantly delayed recovery of psoriasiform dermatitis. We speculate the increased IL-17A in the gastrointestinal tract may be the cause of the less reduction of IL-17A in the skin lesion. The effect of gut microbiota on the distal skin inflammation seems to be achieved by changing the secretion of cytokines, especially IL-17A, to induce systemic inflammation over-activation in psoriasis. These suggest systemic Th17 over-activation or systemic over-secreted IL-17A circulation may be a link between disordered gut microflora and psoriasis development.

In conclusion, multiple evidences we provided here further support the involvement of gut microbiota in psoriatic development. This knowledge provides conceivable promise for developing beneficial supplementary therapeutics for chronic course management of psoriasis. However, further explorations and clinical trials are needed to confirm the validity and safety of FMT in psoriasis.

## Data Availability

The datasets used or analysed during the current study are available from the corresponding author on reasonable request.

## 5. Acknowledgments

This work was supported by Clinical research and translation key project of Sichuan Academy of Medical Sciences & Sichuan Provincial People’s Hospital (No. 2016LZ02), Sichuan Science and Technology Program (No. 2019JDTD0027), and National Natural Science Foundation of China (No. 81573054, 81371729).

## 7. Author Contributions

Chaonan Sun contributed to literature search, operation in experiments, data analysis, statistical analysis, and drafting of the manuscript; Ling Chen contributed to conception of the study, data collection/analysis, literature search, and critical revision of the manuscript; Huan Yang contributed to statistical analysis; Hongjiang Sun contributed to operation in experiments, and statistical analysis; Zhen Xie contributed to data interpretation, and literature search; Bei Zhao contributed to data collection/analysis; Xuemei Jiang contributed to sample collection and data collection; Bi Qin contributed to operation in experiments; Zhu Shen contributed to conception and design of the study, literature search, data collection, statistical analysis, and critical revision of the manuscript.

## Supplementary Material

**Supplementary Fig S1.**
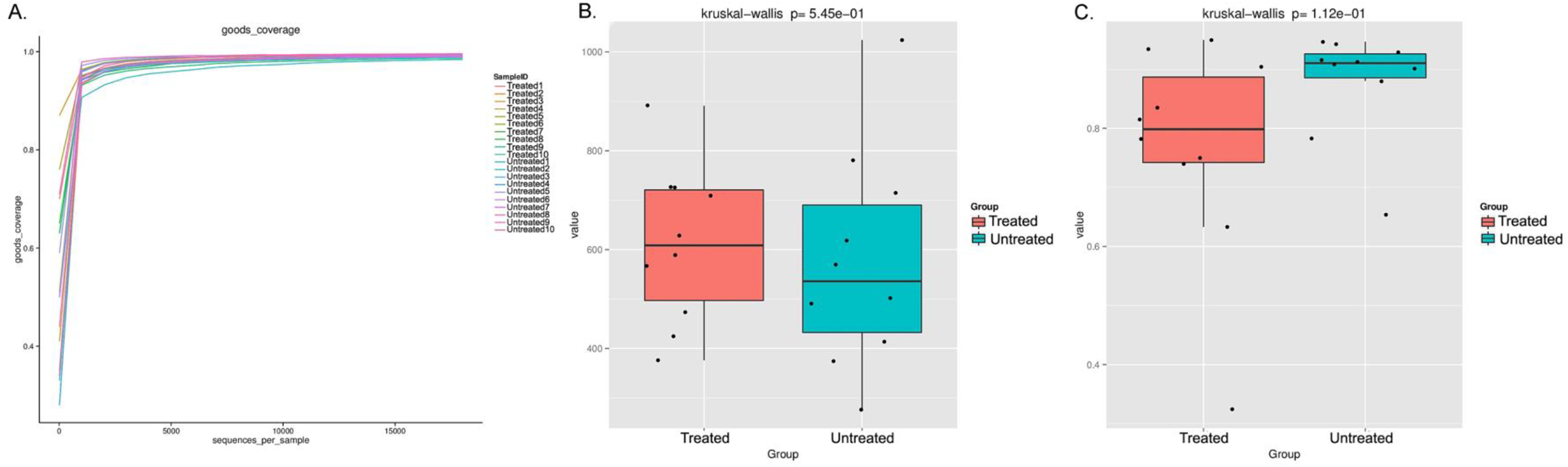
Similar community richness and species diversity in psoriatic patients (un)treated with Acitretin. (A) Obtained goods coverage index rarefaction curves all tended to be plateau, as the reading increases. Box plots with Chao1 (B) and simpon (C) index were depicted.

**Supplementary Fig S2.**
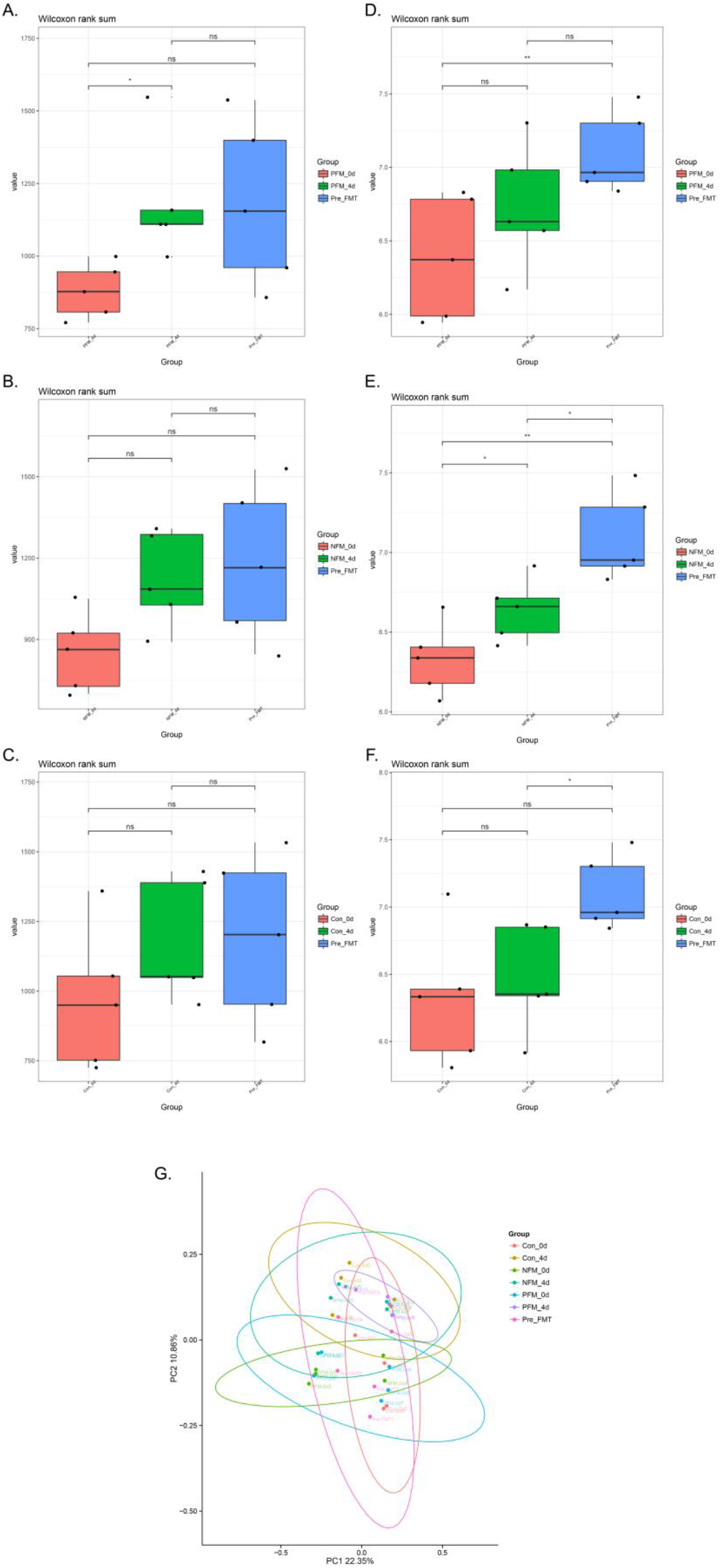
Bacterial Alpha diversity and Beta diversity in all groups of mice. Chao1 index among Pre-FMT, PFM-0d and PFM-4d (A), among Pre-FMT, NFM-0d and NFM-4d (B), among Pre-FMT, CON-0d and CON-4d (C), were depicted with Box plots and analyzed by Wilcoxon rank sum. Shanon index among Pre-FMT, PFM-0d and PFM-4d (D), among Pre-FMT, NFM-0d and NFM-4d (E), among Pre-FMT, CON-0d and CON-4d (F), were depicted with Box plots and analyzed by Wilcoxon rank sum. (G) Principal Coordinates Analysis (PcoA) based on unweighted UniFrac distance showed differential clustering among all group. Solid circles in the same circle represent similar bacterial composition. PC1, principal coordinate 1; PC2, principal coordinate 2. The percentage indicates the contribution of each principal component to the difference. Pre-FMT, before FMT; PFM-0d, at day 0 after FMT of psoriatic fecal sample; NFM-0d, at day 0 after FMT of normal fecal sample; CON-0d, at day 0 after oral gavage of PBS; PFM-4d, at day 4 after FMT of psoriatic fecal sample; NFM-4d, at day 4 after FMT of normal fecal sample; CON-4d, at day 4 after oral gavage of PBS.

## Appendix S1. Main investigation content of gastrointestinal discomfort symptoms in psoriatic patients and general population

1. **In the past five years, your physical condition:**
  ➀ No psoriasis;
  ➁ Psoriasis diagnosed by at least one dermatologist
    If ➀ chose, go directly to **5**.
2. **The total course of your psoriasis:**
  ➀ < 3 months;
  ➁ ≥ 3, and < 6 months;
  ➂ ≥ 6, and < 12 months;
  ➃ ≥ 1, and < 3 years;
  ➄ ≥ 3, and < 5 years;
  ➅ ≥ 5 years
3. **Severity of your psoriasis in the last five years:**
  ➀ Not affecting daily life at all;
  ➁ Slightly affecting daily life;
  ➂ Seriously affecting daily life
4. **Mean area of your psoriasis lesions in the last five years:**
  ➀ < 1 palm;
  ➁ ≥ 1, and < 5 palms;
  ➂ ≥ 5, and < 10 palms;
  ➃ ≥ 10 palms
5. **Full name**:___________(Optional)
6. **Tel**:___________(Optional)
7. **Gender**:
  ➀ male;
  ➁ female
8. **Date of birth**:___________year___________month
9. **Nationality**:
  ➀ Han nationality;
  ➁ Other___________
10. **Marriage status**:
  ➀ unmarried;
  ➁ married;
  ➂ divorced;
  ➃ widowed
11. **Weight (kg)**:___________
12. **Height (CM)**:___________
13. **Long-term residence in recent five years**:___________(province)
14. **Education background:**
  ➀ primary school;
  ➁ junior high school;
  ➂ senior high school or technical secondary school;
  ➃ university or junior college;
  ➄ postgraduate
15. **Monthly income (Yuan)**:
  ➀ none;
  ➁ < 3000;
  ➂ ≥ 3000, and < 5000;
  ➃ ≥ 5000, and < 10000;
  ➄ ≥ 10000

**Do you have the following symptoms:**

**16. Sustained fatigued**:
  ➀ yes;
  ➁ no
**17. Abdominal pain**:
  ➀ no or transient pain;
  ➁ occasional pain affecting part of social activities;
  ➂ prolonged pain affecting most social activities and requiring treatments;
  ➃ severe pain affecting all social activities
**18. Types of abdominal pain**:
  ➀ colic pain;
  ➁ dull pain;
  ➂ tingling pain;
  ➃ cold pain;
  ➄ distending pain;
  ➅ other pain;
  ➆ no pain
**19. Abdominal flatulence**:
  ➀ no or transient abdominal flatulence;
  ➁ occasional abdominal flatulence;
  ➂ frequent and long-term abdominal flatulence;
  ➃ continuous abdominal flatulence, which seriously affects social activities
**20. Borborygmus:**
  ➀ no or transient barborygmus;
  ➁ temporary and occasional barborygmus;
  ➂ frequent and long-term barborygmus;
  ➃ continuous barborygmus, which seriously affects social activities
**21. Gastric acid reflux**:
  ➀ no or transient reflux;
  ➁ occasional reflux;
  ➂ reflux 1-2 times per day, and needs to be treated for relief;
  ➃ reflux several times per day, and anti-acid treatment can only obtain partial relief
**22. Back pain:**
  ➀ no or transient pain;
  ➁ occasional pain to affect part of social activities;
  ➂ prolonged pain to require treatment, and affecting many social activities;
  ➃ severe pain to affect all social activities
**23. Belching**:
  ➀ no or transient belching;
  ➁ occasional belching;
  ➂ frequent and long-term belching, and require treatments for control;
  ➃ persistent belching affecting social activities
**24. Nausea or vomiting**:
  ➀ no nausea;
  ➁ occasionally nausea;
  ➂ frequent and long-term nausea, without vomiting;
  ➃ persistent nausea with vomiting
**25. Passing flatus**:
  ➀ no increased flatus;
  ➁ temporary or occasional increased flatus;
  ➂ increased flatus affecting partial social activities;
  ➃ increased flatus seriously affecting social activities
**26. Urgency of defecation**:
  ➀ normal control;
  ➁ occasional sense of urgency in defecation;
  ➂ frequent sense of urgency in defecation, affecting social activities;
  ➃ fecal incontinence
**27. Constipation**:
  ➀ no constipation;
  ➁ occasional constipation;
  ➂ difficulty in defecation, usually accompanied by feeling of endless defecation;
  ➃ severe constipation, and a treatment is necessary to defecate
**28. Stool frequency**:
  ➀ once a day;
  ➁ 2-3 times per day;
  ➂ more than 3 times per day;
  ➃ once every 2-3 days;
  ➄ once every 4-5 days;
  ➅ once a week or less
**29. Stool color**:
  ➀ yellow brown;
  ➁ green;
  ➂ black;
  ➃ white;
  ➄ no attention
**30. The characteristics of stool:**
  ➀ sross and shaped;
  ➁ thin strip;
  ➂ liquid and shapeless;
  ➃ dry and hard;
  ➄ watery;
  ➅ foamy;
  ➆ no attention

---

**Supplementary Table S1.** The details of the inclusion and exclusion criteria applied to psoriatic patients and general population.

| Participant | Patients | Controls |
| --- | --- | --- |
| Inclusion | 18-60 years<br>Psoriasis diagnosed by at least one dermatologist<br>Traditional Chinese diet | 18-60 years<br>No history of psoriasis and autoimmune disease<br>Traditional Chinese diet |
| Exclusion | Using antibiotics within 1 month<br>Using immunosuppressive agent within 1 month<br>Long-term use of probiotics or prebiotics<br>Long-term consuming yogurt, pickles or cheese<br>Pregnancy<br>A history of acute/chronic gastrointestinal infection, gastrointestinal pathology or gastrointestinal surgery<br>A history of arthritis, enthesitis, or dactylitis<br>Current extreme diet (e.g., vegetarian, parenteral nutrition or macrobiotic diet) | Using antibiotics within 1 month<br>Using immunosuppressive agent within 1 month<br>Long-term use of probiotics or prebiotics<br>Long-term consuming yogurt, pickles or cheese<br>Pregnancy<br>A history of acute/chronic gastrointestinal infection, gastrointestinal pathology or gastrointestinal surgery<br>A history of arthritis, enthesitis, or dactylitis<br>Current extreme diet (e.g., vegetarian, parenteral nutrition or macrobiotic diet) |

**Supplementary Table S2.** The demographic details of psoriatic patients (un)treated with Acitretin.

|  | Untreated patients | Acitretin-treated, over PASI75 | p value |
| --- | --- | --- | --- |
| Sex, n (%) |  |  | 1.0 |
| Male | 5 (50%) | 5 (50%) |  |
| Female | 5 (50%) | 5 (50%) |  |
| Age, mean ± SD | 37.2 ± 14.19 | 36.2 ± 10.58 | 0.87 |
| Disease duration | 4.6 ± 2.37 (year) | 4.78 ± 2.31 (year) | 0.88 |
| PASI score | 15.3 ± 4.11 | 3.6 ± 1.43 | < 0.001 |

PASI75, clinical improvement more than 75%, assessed by PASI score.

**Supplementary Table S3.** The relative abundance of each identified phylum of Untreted group and Treated group.

| OTU | Test-Statistic | P | FDR_P | PRE_mean | POS_mean |
| --- | --- | --- | --- | --- | --- |
| Actinobacteria | 2.643416 | 0.10 | 0.901771 | 0.00922 | 0.003857 |
| Chloroflexi | 2.568276 | 0.11 | 0.901771 | 0.000239 | 0.00038 |
| TA06 | 2.4025 | 0.12 | 0.901771 | 5.43E-06 | 2.71E-05 |
| Lentisphaerae | 2.158589 | 0.14 | 0.901771 | 0.000564 | 7.60E-05 |
| Euryarchaeota | 2.111111 | 0.15 | 0.901771 | 1.09E-05 | 0 |
| Kazan_3B_09 | 2.111111 | 0.15 | 0.901771 | 1.09E-05 | 0 |
| Firmicutes | 1.462857 | 0.23 | 0.973814 | 0.344135 | 0.283062 |
| LCP_89 | 1 | 0.32 | 0.973814 | 1.09E-05 | 0 |
| Elusimicrobia | 1 | 0.32 | 0.973814 | 1.09E-05 | 0 |
| JL_ETNP_Z39 | 1 | 0.32 | 0.973814 | 0 | 5.43E-06 |
| WCHB1_60 | 1 | 0.32 | 0.973814 | 0 | 5.43E-06 |
| Woesearchaeota_(DHVEG_6) | 1 | 0.32 | 0.973814 | 0 | 5.43E-06 |
| Verrucomicrobia | 0.902356 | 0.34 | 0.973814 | 0.000467 | 0.000749 |
| Fusobacteria | 0.632377 | 0.43 | 0.975313 | 0.00427 | 0.02264 |
| Other | 0.600316 | 0.44 | 0.975313 | 0.000157 | 0.000222 |
| Cyanobacteria | 0.521695 | 0.47 | 0.975313 | 0.004655 | 5.43E-05 |
| Candidate_division_OP3 | 0.448148 | 0.50 | 0.975313 | 5.43E-06 | 2.17E-05 |
| Armatimonadetes | 0.448148 | 0.50 | 0.975313 | 5.43E-06 | 1.63E-05 |
| Saccharibacteria | 0.372549 | 0.54 | 0.975313 | 1.09E-05 | 5.43E-06 |
| Hyd24_12 | 0.372549 | 0.54 | 0.975313 | 1.09E-05 | 5.43E-06 |
| Aminicenantes | 0.350985 | 0.55 | 0.975313 | 8.14E-05 | 8.14E-05 |
| Acidobacteria | 0.281057 | 0.60 | 0.99252 | 0.000971 | 0.001042 |
| Bacteroidetes | 0.205714 | 0.65 | 0.99252 | 0.540506 | 0.558209 |
| Chlorobi | 0.146266 | 0.70 | 0.99252 | 0.000244 | 0.000288 |
| Spirochaetae | 0.100496 | 0.75 | 0.99252 | 9.22E-05 | 7.60E-05 |
| Nitrospirae | 0.073077 | 0.79 | 0.99252 | 0.000195 | 0.000239 |
| Gemmatimonadetes | 0.072905 | 0.79 | 0.99252 | 0.00013 | 0.000152 |
| Tenericutes | 0.056763 | 0.81 | 0.99252 | 0.000369 | 0.000195 |
| Latescibacteria | 0.054633 | 0.82 | 0.99252 | 0.000114 | 0.000125 |
| Proteobacteria | 0.051429 | 0.82 | 0.99252 | 0.093213 | 0.128163 |
| Synergistetes | 0.032169 | 0.86 | 0.99252 | 3.26E-05 | 3.26E-05 |
| WD272 | 0.031832 | 0.86 | 0.99252 | 3.80E-05 | 2.71E-05 |
| Planctomycetes | 0.013287 | 0.91 | 1 | 0.000168 | 0.00019 |
| Deferribacteres | 0 | 1.0 | 1 | 2.71E-05 | 2.71E-05 |
| Hydrogenedentes | 0 | 1.0 | 1 | 1.09E-05 | 1.09E-05 |
| Chlamydiae | 0 | 1.0 | 1 | 5.43E-06 | 5.43E-06 |
| Thaumarchaeota | 0 | 1.0 | 1 | 5.43E-06 | 5.43E-06 |

**Supplementary Table S4.** The chao1 and shannon index of mouse fecal microbiota samples.

| Samples | Chao1 | Shannon |
| --- | --- | --- |
| Con-0d1 | 1427.412366 | 7.083762783 |
| Con-0d2 | 693.9056609 | 5.812114353 |
| Con-0d3 | 745.3677374 | 5.924835869 |
| Con-0d4 | 955.8363097 | 6.334937819 |
| Con-0d5 | 1040.601564 | 6.390935075 |
| Con-4d2 | 1404.350522 | 6.869765482 |
| Con-4d5 | 1394.1839 | 6.845933101 |
| Con-4d6 | 1034.892542 | 6.346849332 |
| Con-4d7 | 983.564959 | 5.90159147 |
| Con-4d8 | 1051.095093 | 6.371023789 |
| NFM-0d1 | 722.8400048 | 6.188334112 |
| NFM-0d2 | 736.1812008 | 6.061082875 |
| NFM-0d3 | 874.331424 | 6.402959453 |
| NFM-0d4 | 932.6558627 | 6.351215467 |
| NFM-0d8 | 1029.130211 | 6.668520123 |
| NFM-4d2 | 894.2941169 | 6.415907515 |
| NFM-4d3 | 1282.112356 | 6.918678015 |
| NFM-4d5 | 1265.67648 | 6.665131929 |
| NFM-4d7 | 1071.451454 | 6.725758603 |
| NFM-4d8 | 1046.178209 | 6.482802508 |
| PFM-0d2 | 989.6800906 | 6.784403473 |
| PFM-0d4 | 964.3946533 | 6.375451488 |
| PFM-0d6 | 861.4572118 | 6.828329634 |
| PFM-0d7 | 785.5969888 | 5.952095405 |
| PFM-0d8 | 826.2216702 | 5.979003807 |
| PFM-4d2 | 1570.220641 | 7.293102382 |
| PFM-4d3 | 1163.058397 | 6.996711093 |
| PFM-4d6 | 1106.731235 | 6.552710418 |
| PFM-4d7 | 1141.645119 | 6.636893968 |
| PFM-4d8 | 976.7248754 | 6.156598005 |
| Pre-FMT1 | 829.5077445 | 6.827606612 |
| Pre-FMT2 | 1149.476838 | 6.962579387 |
| Pre-FMT5 | 1534.09003 | 7.510503819 |
| Pre-FMT6 | 1358.612986 | 7.300778446 |
| Pre-FMT7 | 983.5945057 | 6.912840531 |

Pre-FMT, before FMT
PFM-0d, at day 0 after FMT of psoriatic fecal sample
NFM-0d, at day 0 after FMT of normal fecal sample
CON-0d, at day 0 after oral gavage of PBS
PFM-4d, at day 4 after FMT of psoriatic fecal sample
NFM-4d, at day 4 after FMT of normal fecal sample
CON-4d, at day 4 after oral gavage of PBS

**Supplementary Table S5.** The results of Adonis analysis among groups.

|  | **F.Model** | **Pr(>F)** |
| --- | --- | --- |
| Pre-FMT vs PFM-0d vs PFM-4d | 1.6741 | 0.024 |
| Pre-FMT vs NFM-0d vs NFM-4d | 1.4693 | 0.047 |
| Pre-FMT vs PFM-0d vs NFM-0d vs CON-0d vs CON-4d vs NFM-4d vs PFM-4d | 1.6285 | 0.001 |

Pre-FMT, before FMT
PFM-0d, at day 0 after FMT of psoriatic fecal sample
NFM-0d, at day 0 after FMT of normal fecal sample
CON-0d, at day 0 after oral gavage of PBS
PFM-4d, at day 4 after FMT of psoriatic fecal sample
NFM-4d, at day 4 after FMT of normal fecal sample
CON-4d, at day 4 after oral gavage of PBS

